# Assessing the contribution of rare-to-common protein-coding variants to circulating metabolic biomarker levels via 412,394 UK Biobank exome sequences

**DOI:** 10.1101/2021.12.24.21268381

**Authors:** Abhishek Nag, Lawrence Middleton, Ryan S. Dhindsa, Dimitrios Vitsios, Eleanor Wigmore, Erik L. Allman, Anna Reznichenko, Keren Carss, Katherine R. Smith, Quanli Wang, Benjamin Challis, Dirk S. Paul, Andrew R. Harper, Slavé Petrovski

## Abstract

Genome-wide association studies have established the contribution of common and low frequency variants to metabolic biomarkers in the UK Biobank (UKB); however, the role of rare variants remains to be assessed systematically. We evaluated rare coding variants for 198 metabolic biomarkers, including metabolites assayed by Nightingale Health, using exome sequencing in participants from four genetically diverse ancestries in the UKB (N=412,394). Gene-level collapsing analysis – that evaluated a range of genetic architectures – identified a total of 1,303 significant relationships between genes and metabolic biomarkers (p<1×10^−8^), encompassing 207 distinct genes. These include associations between rare non-synonymous variants in *GIGYF1* and glucose and lipid biomarkers, *SYT7* and creatinine, and others, which may provide insights into novel disease biology. Comparing to a previous microarray-based genotyping study in the same cohort, we observed that 40% of gene-biomarker relationships identified in the collapsing analysis were novel. Finally, we applied Gene-SCOUT, a novel tool that utilises the gene-biomarker association statistics from the collapsing analysis to identify genes having similar biomarker fingerprints and thus expand our understanding of gene networks.

## Introduction

Metabolic blood biomarkers represent intermediate or end products of biochemical pathways that can be used to diagnose and monitor human disease. The application of metabolic biomarkers as intermediate traits to dissect the genetic basis of complex human diseases is well-established. Investigating the genetic underpinnings of blood biomarkers can offer novel insights into human disease mechanisms and, in turn, provide potential therapeutic targets. Large-scale genome-wide association studies (GWAS) have so far identified hundreds of genetic loci that regulate blood biomarker and metabolite levels^1–11^; however, difficulty in mapping these loci to causal genes and interpreting functional effects of non-coding variants have stymied the clinical impact for many of these associations^12^.

The UK Biobank (UKB)^13^ is a large population-based resource of ∼500,000 participants with genetic data linked to a diverse set of phenotypic measurements. Genotype data from microarrays and large population-based imputation panels have helped establish the contribution of common and low frequency variants towards blood biomarkers in the UKB^14^. The availability of exome sequences in the same population now allows for the exploration of rare coding variants regulating metabolic blood biomarkers. Associations for rare coding variants have demonstrably greater translational potential given their larger effect sizes^15^ and our ability to more directly interpret their functional impact^16^.

Using exome sequences from 412,394 unrelated participants across multiple genetic ancestries in the UKB, we present findings of variant-level and gene-level (collapsing) association tests for 198 metabolic blood biomarkers. We then introduce a novel tool, Gene-SCOUT, that utilises this rich catalogue of gene-biomarker association statistics to identify genes with similar biomarker fingerprints as a given (target) gene of interest and expand our understanding of gene networks.

## Results

In this study, we analysed 198 metabolic blood biomarkers, including 30 clinical blood biomarkers related to glucose and lipid metabolism, renal and liver function (**Table S1A**), and an additional 168 Nightingale assay blood metabolite measurements related to lipoprotein lipids, fatty acids and their compositions, and various other low-molecular weight metabolites^17^ (**Table S1B**). Most of the metabolic biomarkers pertain to lipid metabolism (77%) and correlate highly with each other (**Figure 2a**). Many metabolic biomarkers also demonstrate strong associations with clinical traits documented in the UKB (**Figure 2b**).

We first conducted a single variant analysis between the non-synonymous coding variants (N=2,043,019 for the European ancestry subset) and the 198 metabolic biomarkers (**Figure 1**). Excluding the MHC region, 19,351 significant variant-biomarker associations (p<1×10^−8^) were identified in the European subset of UK Biobank, which mapped to 12,217 significant relationships between genes and biomarkers (**Tables S2A, S2B**). Pruning variants in linkage disequilibrium (r^2^ threshold of 0.5) resulted in 9,738 significant gene-biomarker relationships. Notably, 243 distinct PTVs accounted for 1,366 significant associations, of which 602 (44%) were attributable to rare PTVs (MAF<0.1%) with large effect sizes (>0.5 SD) (**Figure 4a**) (**Tables S3A, S3B**). We identified 28 PTVs with MAF as low as 0.001% that achieved significance in the ExWAS: these include associations relating to several well-established and biologically plausible relationships such as *GOT1* and aspartate aminotransferase, *CST3* and cystatin C, *APOB* and cholesterol biomarkers, *ALPL* and alkaline phosphatase (**Table S3A**). Among other PTV findings that may provide new insights into important biology, a rare frameshift variant (MAF=0.03%) in *PLIN1 – a* gene known to cause familial lipodystrophy^18^ – was associated with HDL-cholesterol (beta=0.40 [0.27,0.53], p=1.6×10^−9^), and a rare splice variant (MAF=0.09%) in *TNFRSF10B* – loss of which has been reported to promote survival of virus-infected liver cells^19^ – was associated with gamma glutamlytransferase (beta=0.21 [0.14,0.28], p=3.8×10^−9^) (**Table S3A**).

**Figure 1:**
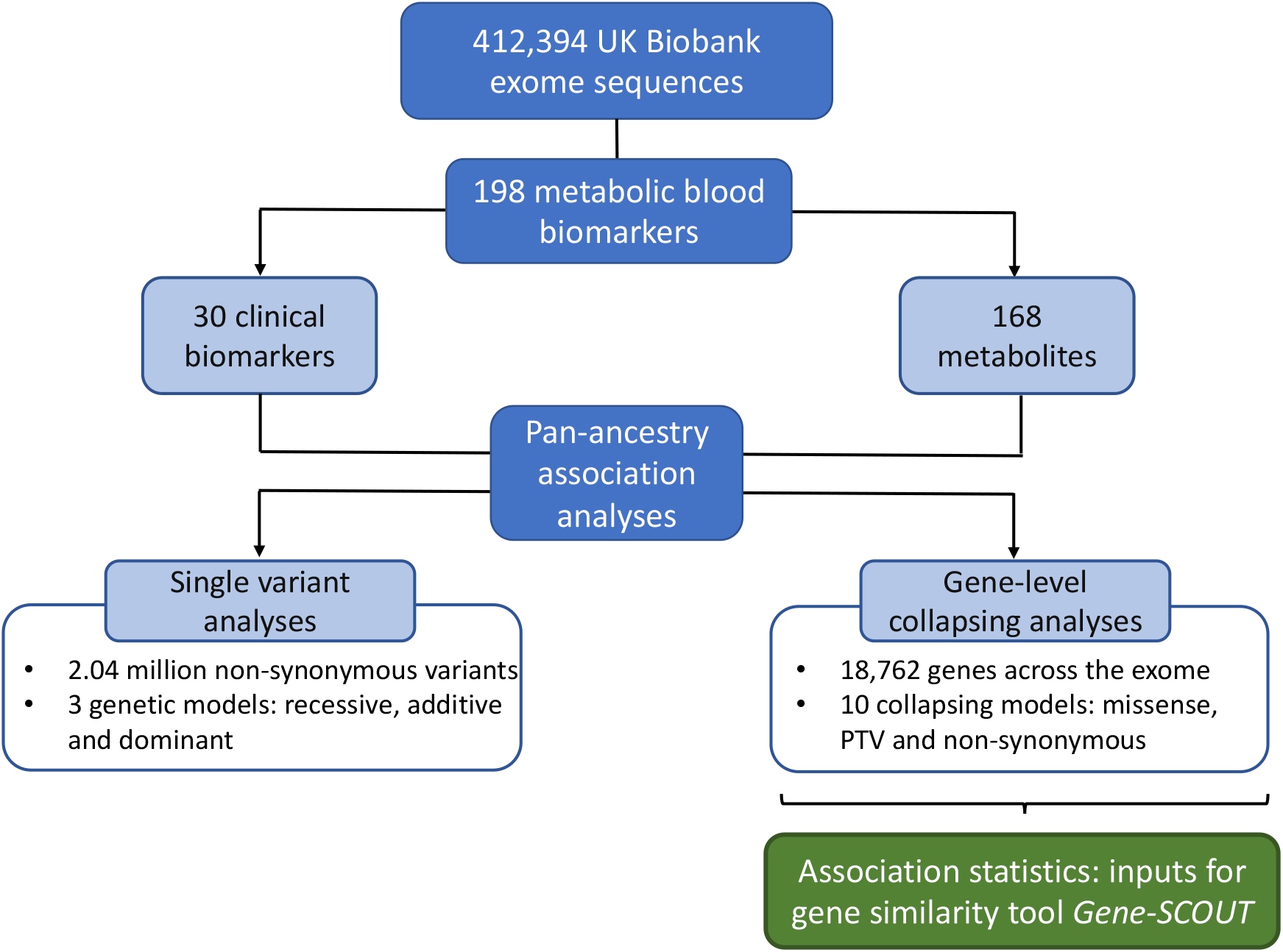
A schematic of the association analyses that were conducted for the metabolic blood biomarkers using the UK Biobank exome sequences. The UK Biobank exome sequences were used to conduct single variant (under 3 genetic models) and gene-level (under 10 collapsing models) association analyses for the clinical blood biomarkers (N=30) and blood metabolite measurements (N=168). The gene-level association statistics for these metabolic biomarkers were used as inputs for the gene similarity tool *Gene-SCOUT*.

**Figure 2:**
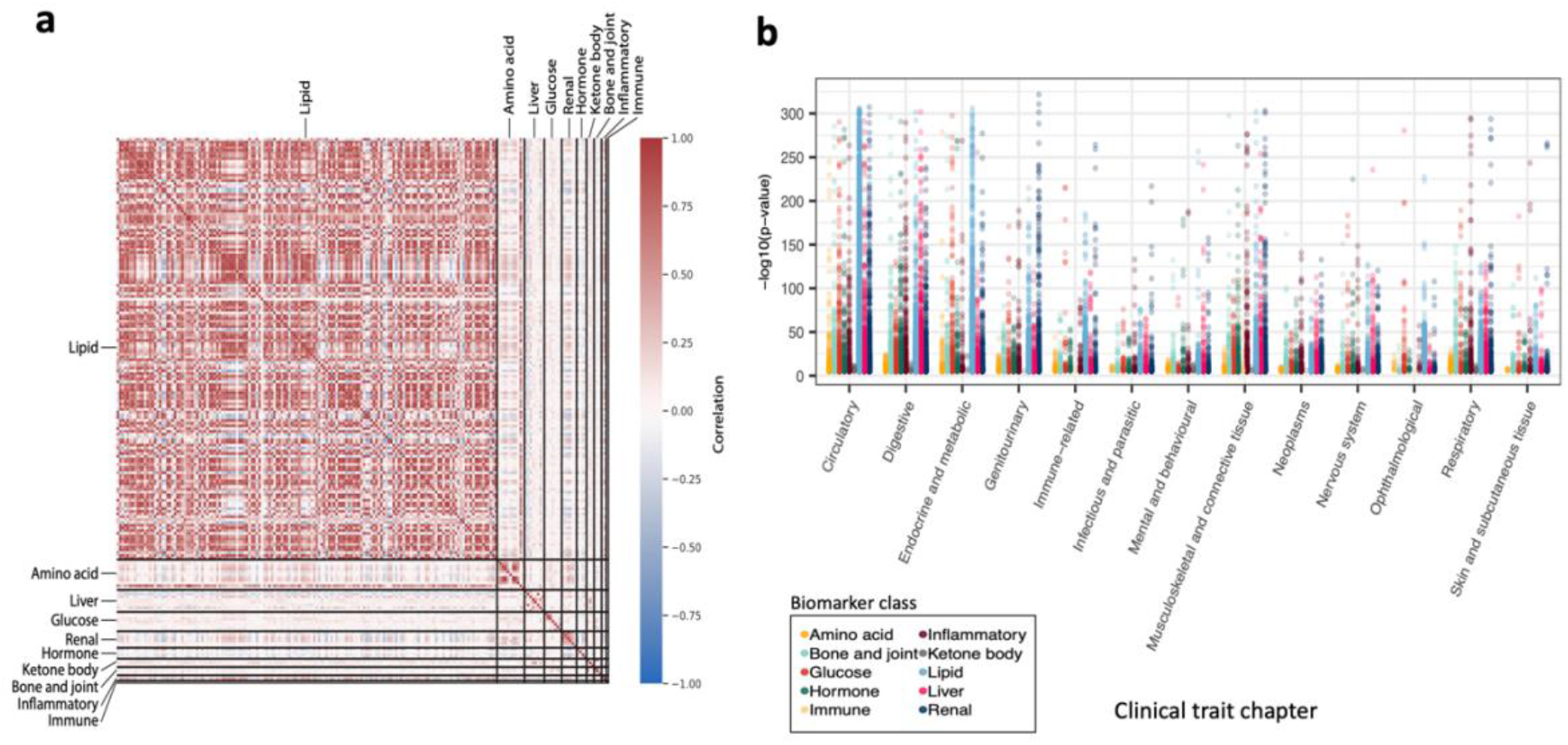
Characteristics of metabolic blood biomarkers analysed in this study. The 198 metabolic blood biomarkers analysed in this study were grouped into the following 10 biological classes: lipid, amino acid, liver, glucose, renal, hormone, ketone body, bone and joint, inflammatory, and immune. (a) The plot demonstrates that metabolic biomarkers belonging to the same biological class are correlated with each other. (b) Strong associations (plotted on the Y-axis) were observed between the metabolic biomarkers and 15,719 clinical traits (grouped by chapter) documented in the UKB^20^.

Next, we performed a gene-level collapsing analysis that tests the aggregate effect of rare functional variants in each gene. We employed 10 different models to capture a diverse range of genetic architectures (**Methods**). In the analysis involving individuals of European ancestry alone, we identified 1,303 significant relationships between genes and metabolic biomarkers (p<1×10^−8^) (**Tables S4A, S4B; Figures 3a, 3b**). Most (68%, 880/1,303) gene-biomarker relationships detected via the collapsing analysis were captured through models that exclusively focused on PTV classes (“ptv” and “ptv5pcnt”), while the remaining 32% were attributable to models that incorporated missense variants. We detected more significant associations using our “ptv” and “ptv5pcnt” models than a prior study^21^ that also performed gene-level collapsing analysis using the UKB exome sequence data, albeit with a different analytical framework. For instance, associations between PTVs in 12 genes and HbA1c that we detected were not reported in the other study: this includes the glucose metabolism genes *HK1* and *G6PC2* (**Figure S1)**. We also extended our gene-level collapsing analysis to include all ancestral groups in the UKB (**Methods**). This detected an additional 51 significant gene-biomarker relationships (**Tables S5A, S5B**). For the gene-biomarker relationships that were significant only in the pan-ancestry analysis, we did not observe a significant difference in the estimated effect size between the European-only and the pan-ancestry analyses (p=0.83), suggesting that increased statistical power rather than ancestry-specific effects is the more likely reason why these associations were identified in the pan-ancestry analysis. One such association detected exclusively in the pan-ancestry analysis was between recessive carriers of nonsynonymous variants in the membrane transport gene *SYT7* and blood creatinine levels (number of QV carriers=5, beta=2.17 [1.46,2.87], p=1.6×10^−9^). With 3 of the 5 carriers observed in the South Asian and African ancestry participants, the pan-ancestry analysis facilitated detection of this association, which was not study-wide significant in the European subset (number of QV carriers=2, beta=1.17 [0.06,2.28], p=0.04). Remarkably consistent with the biomarker findings, recessive carriers of *SYT7* PTVs demonstrate a increased risk of glomerular disease in the pan-ancestry analysis (OR=92.1 [12.1,713.2], p=2.6×10^−5^), but the clinical association on its own is not yet study-wide significant.

**Figure 3:**
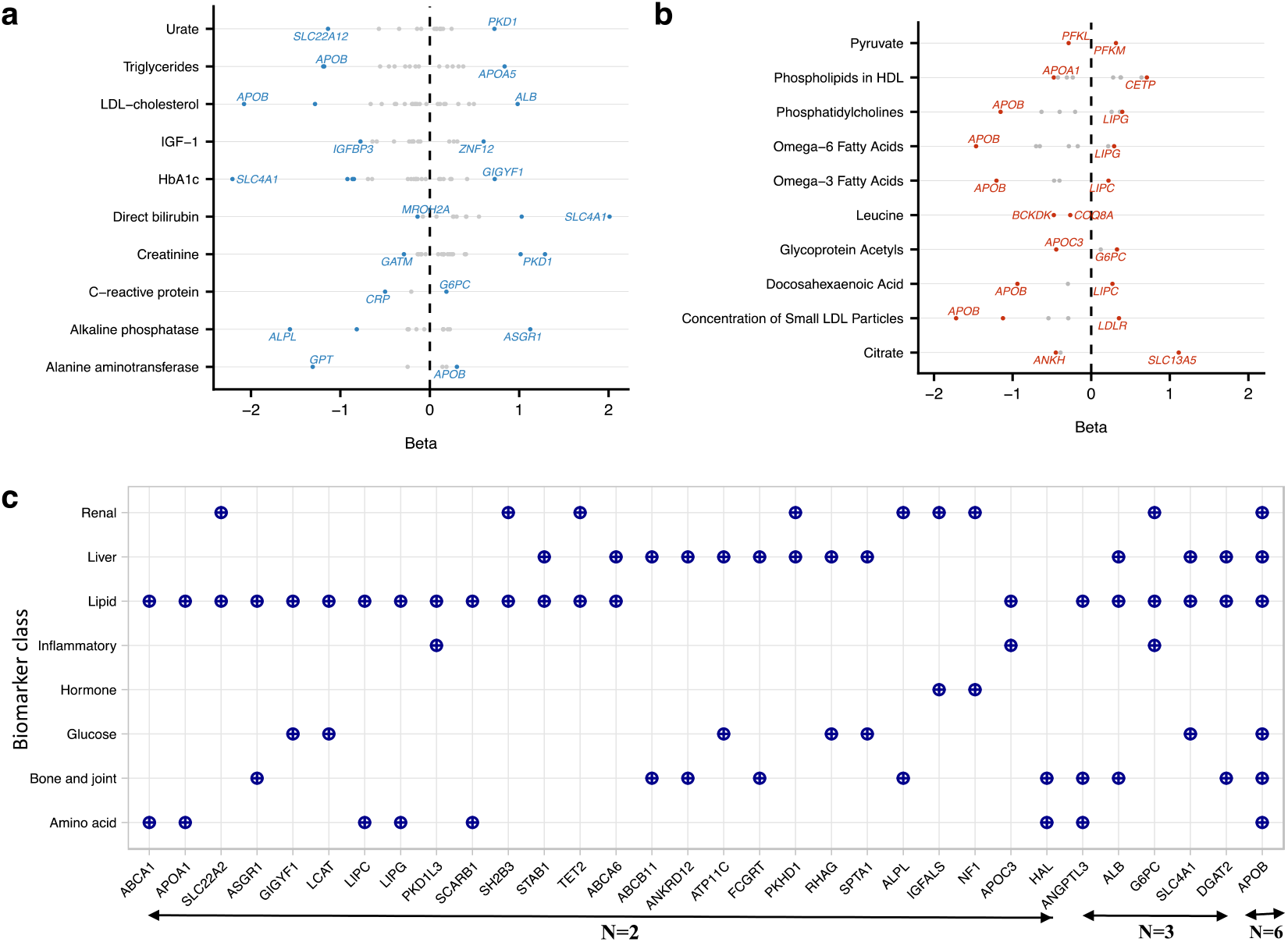
Significant relationships between genes and metabolic blood biomarkers identified in the collapsing analysis. (a, b) Significant gene relationships (p<1×10^−8^) identified for select clinical biomarkers and metabolites in the collapsing analysis have been shown. The genes with the highest absolute effect sizes for each have been labelled. (c) The plot lists the 32 genes that were significantly associated (p<1×10^−8^) with metabolic biomarkers across two or more biological classes in the collapsing analysis. For each such gene, the corresponding biological classes have been indicated.

The significant gene-level relationships from the collapsing analyses encompassed 207 distinct genes, of which 32 were associated with biomarkers across different biological classes (**Figure 3c**). This includes *GIGYF1*, a tyrosine kinase receptor signalling protein, in which rare PTVs were associated with biomarkers of glucose [glucose (beta=0.59 [0.42,0.76], p=7.9×10^−12^) and HbA1c (beta=0.73 [0.57,0.88], p=4.5×10^−20^)] and cholesterol metabolism [total cholesterol (beta=-0.66 [-0.82,-0.50], p=2.0×10^−15^), LDL-cholesterol (beta=-0.61 [-0.78,-0.45], p=3.4×10^−13^) and apolipoprotein B (beta=-0.60 [-0.77,-0.44], p=1.3×10^−12^)]. Additionally, among clinical traits documented in the UKB^20^, significant associations were observed for rare PTVs in *GIGYF1* with the risk of hypothyroidism (OR=4.2 [2.7,6.6], p=7.1×10^−9^) and type 2 diabetes (OR=4.0 [2.7,5.8], p=1.0×10^−10^). Since hypothyroidism is known to raise LDL-cholesterol levels, we subsequently tested the *GIGYF1*–LDL-cholesterol association adjusted for a diagnosis of hypothyroidism. The signal between *GIGYF1* PTVs and LDL-cholesterol (adjusted for the effect of statins) remained significant upon adjusting for hypothyroidism (beta=-0.55 [-0.71,-0.38]; p=6.2×10^−11^), suggesting that the *GIGYF1* locus likely influences cholesterol levels independent of solely thyroid hormone-mediated pathways. Thus, by leveraging information from over 400,000 UKB exomes, our study provides a more comprehensive picture regarding *GIGYF1*’s biomarker fingerprint and associated clinical traits, expanding on previously reported common^7^ and rare variant associations^21,22^ at this locus.

We observed that adjusting biomarkers for medications that influence their levels can also improve detection of associations: 31/84 (37%) significant gene-biomarker relationships for apolipoprotein B, LDL-cholesterol, total cholesterol, and urate from the collapsing analysis were detected only after we adjusted their values for commonly prescribed medications (**Table S4A**). This includes association between putatively damaging missense variants and PTVs in *HMGCR* (“flexdmg” model) and LDL-cholesterol (medication-adjusted: beta=-0.19 and p=1.7×10^−11^; medication-unadjusted: beta=-0.15 and p=6.1×10^−8^), which validates the value of medication adjustment to untangle the effects of therapeutic intervention vs natural aberration of *HMGCR*. Moreover, for gene-biomarker relationships that were significantly associated in both the medication-unadjusted and the medication-adjusted analyses (N=52), the absolute effect sizes were observably higher in the latter (**Figure S2**), but the difference was not statistically significant in the current sample (Mann Whitney p=0.28).

### Gene-level collapsing analysis: capturing allelic series

We observed that 17% (215/1,303) of significant relationships between genes and metabolic biomarkers from the collapsing analysis did not achieve significance in the respective variant-level ExWAS (**Table S6**). Next, we also compared the gene-biomarker relationships that achieved significance in the collapsing analysis (**Tables S4A, S7**) and the microarray-based GWAS^14^ (as per a less stringent significance threshold: p<1×10^−7^) for the 32 biomarkers (28 blood and 4 urinary biomarkers) analysed in both studies. Of the significant gene-biomarker relationships identified in the collapsing analysis, 40% (142/357) were not detected in the microarray-based GWAS (**Table S8**). These include associations for well-known drug target genes such as *HMGCR* (with LDL-cholesterol) and *PPARG* (with HDL-cholesterol). Furthermore, the effect size estimates were significantly higher in the collapsing analysis than in microarray-based GWAS for 215 gene-biomarker relationships detected via both approaches (Mann-Whitney p=8.0×10^−6^) (**Table S9; Figure 4b**). One likely explanation for this is that by testing aggregate effects of rare putative functional variants in a gene, associations arising from collapsing analysis are enriched for larger effects (**Figure 4c**). Collectively, these results highlight that application of a gene-based rare variant collapsing analysis to large-scale exome sequencing can increase power to capture associations that are driven by an allelic series, and thus expand our understanding of the genetic architecture of traits, especially where a lot of success has already been achieved through traditional microarray-based GWAS.

**Figure 4:**
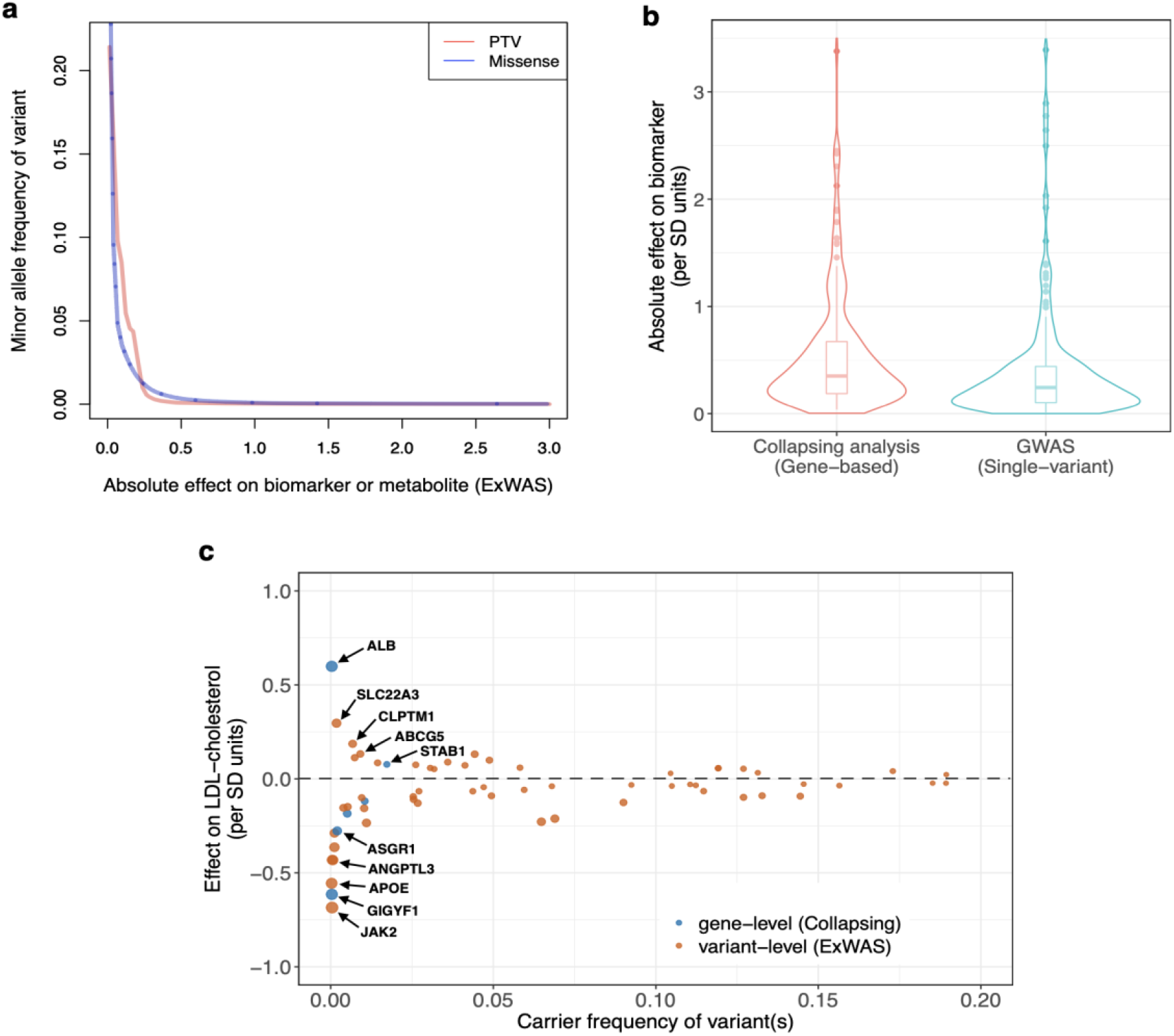
Effects of coding variants on metabolic blood biomarkers. (a) Absolute effect sizes for missense variants and PTVs significantly associated (p<1×10^−8^) with metabolic biomarkers in the single variant analysis (ExWAS) as a function of their minor allele frequency (in cases where a missense variant or a PTV was significantly associated with more than one biomarker, the association with the highest absolute effect size was selected). (b) The effect sizes estimated in the gene-based collapsing analysis and the Sinnott-Armstrong *et al* microarray-based GWAS^14^ were compared for the gene-biomarker relationships that were significantly associated in both (N=215). For each significant gene-biomarker relationship, the collapsing model (from the collapsing analysis) and the individual variant (from the microarray-based GWAS) with the highest absolute effect sizes were selected. The effect sizes estimated in the collapsing analysis were significantly higher than that in the GWAS (Mann-Whitney p=8.0×10^−6^). (c) Comparing effect sizes for individual variants and aggregate of rare variants (in a gene) that were significantly associated (p<1×10^−8^) with LDL-cholesterol. Some examples of genes significantly associated with LDL-cholesterol have been highlighted. The Y-axis has been capped at 1 SD units for visual clarity.

*SLC4A1*, which encodes a chloride/bicarbonate anion exchange protein in the red cell membrane, represents one such gene for which multiple signals were detected in the gene-level collapsing analysis but not in the ExWAS. We observed 32 carriers for 28 distinct *SLC4A1* PTVs, of which 25 (89%) were private (i.e., observed in a single carrier) (**Figure S3**). Overall, *SLC4A1* PTVs were significantly associated with a strong reduction in HbA1c (beta=-2.2 [-2.6,- 1.8], p=1.4×10^−25^) and LDL-cholesterol (beta=-1.0 [-1.4,-0.7], p=8.0×10^−9^), while also showing strong increases in total bilirubin (beta=1.7 [1.3,2.0], p=1.1×10^−22^) and direct bilirubin (beta=2.0 [1.7,2.4], p=1.8×10^−28^). Among clinical phenotypes, *SLC4A1* PTVs are significantly associated with disorders of reduced red cell membrane stability such as hereditary spherocytosis and hereditary haemolytic anaemia, but not with any phenotype related to glucose or lipid metabolism (p<1×10^−5^). Similarly, in ClinVar, several missense and loss-of-function mutations in this gene are reported as pathogenic for hereditary spherocytosis. Therefore, we further tested the *SLC4A1*–biomarker associations after adjusting for the diagnosis of hereditary spherocytosis or hereditary haemolytic anaemia and relevant blood cell indices, including red cell distribution width (RDW) and mean corpuscular haemoglobin concentration (MCHC). The gene-based *SLC4A1* PTV signals remained significant in the adjusted analyses (**Table S10**). Although PTVs in this gene may be independently associated with biomarkers of glucose, lipid and bilirubin metabolism, we cannot rule out the possibility of under-reporting of hereditary spherocytosis and hereditary haemolytic anaemia in the UKB that explains these observations. The *SLC4A1* enigma is consistent with previous reports of other red blood cell loci that have also been significantly associated with HbA1c^23^.

### Gene-SCOUT: estimating gene similarity based on cohort statistics from collapsing analysis

We considered the opportunity to leverage this new and rich catalogue of gene-level association statistics from the collapsing analysis to determine genes with similar biomarker fingerprints. To achieve this, we developed a gene similarity tool ‘Gene-SCOUT’^24^, that solely uses the gene-level collapsing analysis statistics across the studied biomarkers to identify genes with the most comparable biomarker genetic associations as a given gene of interest. No other information is used in constructing the gene similarity scores. Since this tool estimates gene similarity for an index gene by selecting features based on the significance cut-off of p<1×10^−5^, gene neighbours could not be determined for genes that did not achieve association p<1×10^−5^ with any biomarker feature. Accordingly, for our feature set comprising of 198 biomarkers, we were able to determine gene similarity for 3% (536/18,762) of human protein-coding genes. To illustrate Gene-SCOUT’s application, we selected the 24 genes that were significantly associated (p<1×10^−8^) with LDL-cholesterol in the collapsing analysis. We used each gene in this set as a seed gene to construct a network figure that demonstrates their respective gene neighbours (**Figure 5**). Using *APOB* as an example, we observe that genes with the most comparable biomarker fingerprint as *APOB* include: *ABCA1, ACVR1, APOC3, ANGPTL3, ASGR1, ASXL1, BTNL9, GIGYF1, HIST2H2BE, HMGCR, NPC1L1, PCSK9, PDE3B, PKD1L3, RRBP1, SLC4A1, TM6SF2* and *ZNF229*. For some of these genes (e.g., *ZNF229, ACVR1*), the links with lipid metabolism appear to be novel, in addition to the recently described relationships for *GIGYF1*^21,22^. Inhibition of *APOB*, such as through mipomersen, is known to be clinically effective in reducing blood cholesterol levels. Remarkably, 5 (namely, *APOC3, ANGPTL3, HMGCR, NPC1L1* and *PCSK9*) of the 18 genes (28%) determined to have similar cohort-level genetic associations for biomarkers as *APOB* are also targets of lipid-lowering drugs that are already approved or in various stages of development (https://www.fda.gov/drugs).

**Figure 5:**
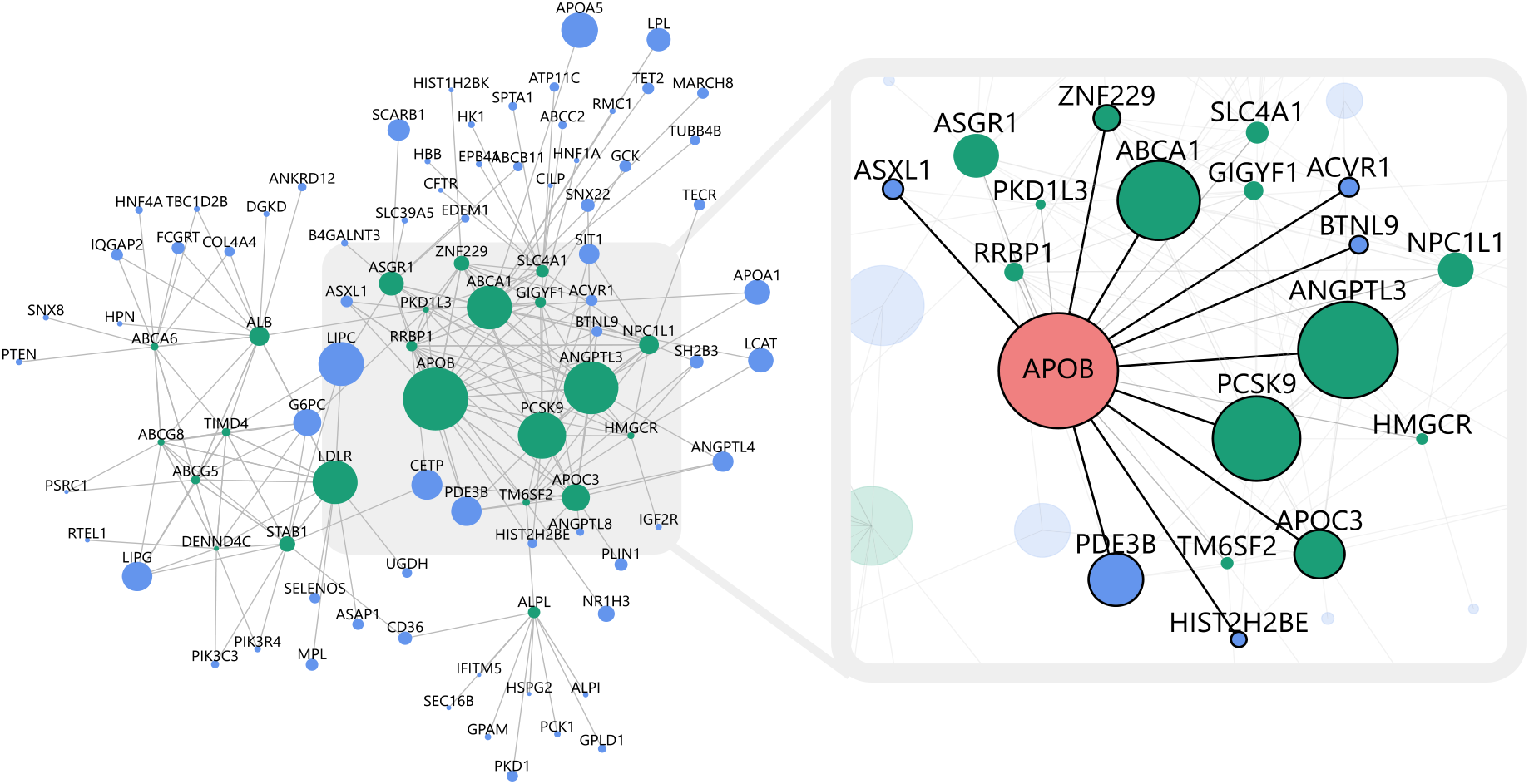
**Network figure demonstrating the gene neighbours i.e., genes with most similar biomarker genetic signals, as the set of genes that were significantly associated with LDL-choleserol in the collapsing analysis** The 24 genes that were significantly associated (p<1×10^−8^) with LDL-cholesterol in the collapsing analysis were used as seed genes (green nodes) to construct a network figure demonstrating respective gene neighbours (edges). Non-seed genes are represented using blue nodes. The size of a gene node corresponds to the number of features (of total 198) that the gene is associated with at p<1×10^−5^. The inset demonstrates the genes with most similar biomarker signature as *APOB* – these include the ten closest genes for *APOB* as the seed gene (black edges) and other seed genes that have *APOB* among their ten closest genes (grey edges).

## Discussion

We used the 454,796 UK Biobank exome sequences to explore the contribution of private-to-rare-to-common protein-coding variation for 30 clinical biomarkers and 168 metabolite measurements. By adopting variant- and gene-level analysis frameworks and assessing the full allelic frequency spectrum, we have expanded our understanding of the genetic architecture of metabolic biomarkers that have previously been studied through microarray data. The finding that 17% of gene-biomarker relationships detected in the gene-level collapsing analysis were not identified in the single variant analysis demonstrates the power of testing an aggregate effect of rare variants in a gene encompassing a range of genetic architectures. We also illustrated how adjusting biomarker values for commonly prescribed medications can improve signal detection.

There are several strengths of our study that might have implications for identifying or validating drug targets. First, by virtue of focusing on coding variants, the observed associations could provide a more causal link between a gene and a blood biomarker^25–28^. Moreover, association signals emerging from collapsing analysis are driven by an aggregate effect of multiple rare variants (allelic series) that tend to be less impacted by local LD structure. This contrasts with associations identified in microarray-based GWAS that often map to non-coding regions of the genome or to regions of extensive LD, making it more challenging to pinpoint the underlying causal variants.

Associations involving putative functional variants can also indicate the desired modulation of the target gene e.g., upregulation or downregulation of the target gene product, required to mitigate the risk of the disease related to the associated biomarker. For instance, we observed a total of 182 associations for rare (MAF<0.1%) PTVs with the 30 blood biomarkers, which is >3-fold more than the 53 conditionally independent PTV associations (for the same set of blood biomarkers) reported in the microarray-based analysis^14^.

We also introduce a novel tool (Gene-SCOUT) that utilises all the gene-level collapsing analysis statistics across the 198 studied biomarkers to estimate a ‘similarity’ metric between genes. With the aid of specific examples, we were able to demonstrate that this approach can successfully identify genes with similar biomarker fingerprints.

While there are certain advantages of using blood biomarkers to dissect the genetics of complex human diseases, including greater statistical power offered by quantitative traits and better insights into biological pathways underlying associations, further work is necessary to establish the causal relationship between genetic loci identified using biomarkers or metabolites and the related disease(s). For instance, we observed associations between certain biomarkers and variants in genes that encode them (e.g., *ALB* with albumin, and *CST3* with cystatin C) – although such associations serve as excellent positive controls that demonstrate the robustness of our analysis framework, they may not offer novel insights into disease pathophysiology.

Using the largest collection of exome sequences linked to a diverse set of circulating metabolic biomarkers, we demonstrate the value of this resource to enhance our understanding of human diseases, and potentially, provide novel therapeutic targets focused on mimicking natural human genetic discoveries. Our study also strongly supports the use of a gene-based collapsing framework to uncover gene-biomarker relationships that are driven by an aggregate effect of multiple rare, non-synonymous variants.

## Methods

### UK Biobank (UKB) Resource

The UKB resource^13^ is a prospective cohort study of ∼500,000 individuals from across the United Kingdom, aged between 40 and 69 years. The average age at recruitment for the sequenced participants was 56.5 years and 54% of the sequenced cohort are females. Participant data, obtained through questionnaires and assessment visits, include health records that are periodically updated by the UKB, self-report survey information, linkage to death and cancer registries, urine and blood biomarkers, imaging data, accelerometer data and various other phenotypic endpoints^13^. All study participants provided informed consent. For this study, data from the UKB resource was accessed under the application number 26041.

### Metabolic blood biomarkers

Routine clinical blood biomarkers related to glucose and lipid metabolism, renal and liver function, among others (N=30), were measured in the majority of the ∼500,000 UKB participants (**Table S1A**). Additionally, 168 blood metabolites, including lipoprotein lipids, fatty acids and their compositions, and various low-molecular weight metabolites, were profiled in a subset of ∼120,000 UKB participants by Nightingale Health using nuclear magnetic resonance spectroscopy^17^ (**Table S1B**). Samples with a ‘quality control (QC) flag’ for the blood metabolites were excluded. In total, we analysed 198 metabolic blood measures: 30 clinical biomarkers and 168 metabolites. We applied rank-based inverse-normal transformation to the measurements prior to performing association analyses.

For four blood biomarkers (LDL-cholesterol, total cholesterol, apolipoprotein B and urate) we adjusted for the effect of commonly prescribed medications known to influence their levels. For LDL-cholesterol, total cholesterol and apolipoprotein B, we adjusted for the effect of statins based on their ‘statin adjustment factors’, previously estimated in the UKB as 0.684, 0.749 and 0.719, respectively^14^. Similarly, we adjusted urate for the effect of allopurinol based on an ‘allopurinol adjustment factor (0.810)’, calculated using an approach identical to that described for statins^14^.

### Whole-exome sequencing and bioinformatics pipeline

Whole-exome sequences for 454,988 UKB participants were generated at the Regeneron Genetics Center as part of a pre-competitive data generation collaboration between AbbVie, Alnylam Pharmaceuticals, AstraZeneca, Biogen, Bristol-Myers Squibb, Pfizer, Regeneron and Takeda^29^. The exome sequencing procedure and the relevant QC steps have been detailed previously in Szustakowski *et al* (2021)^29^ and Wang *et al* (2021)^20^. The FASTQ sequences that were made available were first aligned, following which, single nucleotide variants (SNVs) and small indels were called using Illumina’s DRAGEN Bio-IT Platform Germline Pipeline v3.0.7 on the Amazon Web Services cloud compute platform available at AstraZeneca’s Centre for Genomics Research. SNPEff v4.3^30^ was used to annotate the ‘most damaging effect’ predicted for each protein coding variant. In addition, we used certain other bioinformatic tools such as missense tolerance ratio (MTR) scores^31^ to identify regions of protein coding genes under constraint for missense variants, and REVEL^32^ to prioritise coding variants based on their predicted deleteriousness. Further details on how these tools were applied to the UKB exome sequencing dataset have been previously described^20^.

### Selection of UKB samples for the association analyses

Prior to performing the association analyses, we excluded samples from the available UKB exome sequencing dataset (N=454,796) based on the following QC measures^20^ (**Figure S4**):

i. *DNA contamination*: VerifyBAMID freemix (measure of DNA contamination) >4%.
ii. *Coverage depth*: ≥10x for <94.5% of the consensus coding sequence (CCDS release 22).
iii. *Relatedness*: 2^nd^-degree relatives or closer (equivalent to kinship coefficient>0.0884), as estimated using the --kinship function in KING v2.2.2^33^.

Additionally, to perform analyses accounting for differing genetic ancestry, we assigned samples to one of the four major ancestral groups (minimum 1,000 participants): European (N=394,695), South Asian (N=8,078), East Asian (N=2,209) and African (N=7,412). This was done by excluding participants: (i) with predicted genetic ancestry <0.99 (for European ancestry) or <0.95 (for the remaining ancestries), as estimated using PEDDY v0.4.2; or (ii) lying outside four standard deviations for the top four principal components for each of the genetic ancestry collections.

### Association analysis for metabolic blood biomarkers

A number of stringent variant-level QC steps, detailed previously^20^, were applied to select variant calls with highest confidence for association testing. Briefly, the variant-level QC criteria included coverage depth, genotype and mapping quality scores, DRAGEN variant status, read position rank sum score (RPRS), mapping quality rank sum score (MQRS), alternate allele read proportion for heterozygous calls, proportion of samples failing any of these QC criteria, and gnomAD-related filters.

Association testing between the metabolic blood biomarkers and the variants in the exome sequencing dataset was conducted using two complementary analytical approaches (**Figure 1**):

i. Single variant exome-wide association study (ExWAS)
ii. Gene-level collapsing analysis

We conducted the association analyses separately in the European ancestry participants as this comprised the single largest ancestral group in this resource and for all four ancestries combined (‘pan-ancestry’ analysis).

### Single variant exome-wide association study (ExWAS)

In the single-variant analysis (hereafter referred to as ‘ExWAS’), variants that passed the QC steps were filtered further to include those that had a minimum of six carriers (equivalent to MAF>0.0008% in the European ancestry subset). We additionally excluded variants that had one of the following annotations as their most damaging effect as per SNPEff: *3_prime_UTR, 5_prime_UTR, initiator_codon_variant, non_coding_transcript_exon_variant*, and *synonymous_variant*. The remaining non-synonymous coding variants (N=2,043,019 in the European ancestry subset) were used to perform the ExWAS.

The ExWAS was conducted by fitting a linear regression model adjusted for age, sex and BMI (for blood metabolites only), using the tool PEACOK that was developed as a modification of the R package PHESANT^34^. For the pan-ancestry analysis, we additionally included the categorical ancestral group and top five ancestry principal components as covariates. For each of the 198 biomarkers, three different genetic models were evaluated in the ExWAS: (i) genotypic (AA vs AB vs BB), (ii) dominant (AA+AB vs BB), and (iii) recessive (AA vs AB+BB), where A and B denote the reference and alternative alleles, respectively. A significance cut-off of p<1×10^−8^ was adopted for the ExWAS^35^.

### Gene-level collapsing analysis

In order to boost power to detect associations for rare variants (including private mutations) having the same direction of effect, we adopted a collapsing framework to test the aggregate effect of rare functional variants in a gene. Overall, 10 different collapsing models (9 dominant and one recessive) were implemented per gene to evaluate a range of genetic architectures. Additionally, a synonymous collapsing model was used for the purpose of establishing an empirical negative control^20^.

As outlined in **Table S11**, the criteria for qualifying variants (QVs)^36^ for the collapsing models were based on the following parameters: type of variant (missense, non-synonymous or PTV), minor allele frequency, *in silico* deleteriousness predictors (REVEL and MTR), and type of genetic model (dominant or recessive). The following variant annotations were used to define PTVs: *exon_loss_variant, frameshift_variant, start_lost, stop_gained, stop_lost, splice_acceptor_variant, splice_donor_variant, gene_fusion, bidirectional_gene_fusion, rare_amino_acid_variant* and *transcript_ablation*. Hemizygous genotypes for the X chromosome also qualified for the recessive model.

For a given collapsing model, the effect of QVs in each gene (N=18,762) was calculated as the difference in the mean of a blood biomarker between carriers and non-carriers of the QVs, using a linear regression model in PEACOCK. Covariates used in the linear regression model were identical to that described for the ExWAS.

A significance cut-off of p<1×10^−8^ was set for the collapsing analysis based on the observed p-value distribution for the synonymous model and an n-of-1 permutation, as described previously^20^.

### Association analysis of clinical phenotypes documented in the UKB

We harmonized and union mapped the clinical phenotypes available in the UKB, as previously described^20^. Phenome-wide collapsing analysis for 15,719 clinical phenotypes was performed for the 11 collapsing models, as described in our previously published study^20^. We queried the results of this analysis for genes of interest that emerged from the analysis of the metabolic biomarkers.

Additionally, we also performed an association analysis between the each of the 198 metabolic biomarkers and the clinical phenotypes using a linear regression model adjusted for age and sex.

### Comparison of results from collapsing analyses to microarray-based genome-wide association study

We explored the hypothesis that the application of a collapsing framework – that tests the aggregate effect of rare functional variants in a gene identified using exome sequencing – detected gene-biomarker relationships that were previously not identified in microarray-based studies. In order to do that, we compared our findings with the results from a recent study^14^ that conducted single variant association analysis (GWAS) for clinical biomarkers in the UKB using microarray data, including directly genotyped coding variants. Besides the 28/30 clinical blood biomarkers that we studied, seven other biomarkers (mainly, urine-related) were analysed in the GWAS. These seven biomarkers comprised of four urinary biomarkers that were directly measured in the UKB and an additional three derived measurements. For the purpose of comparing findings, we additionally performed gene-level collapsing analysis for the four urinary biomarkers for which data were directly available in the UKB (i.e. ‘sodium in urine’, ‘potassium in urine’, ‘microalbumin in urine’, and ‘creatinine (enzymatic) in urine’). To be consistent with the microarray-based GWAS, we used the statin-adjusted values for LDL-cholesterol, total cholesterol, and apolipoprotein B, and the medication-unadjusted values for the remaining biomarkers. Thereafter, for the set of 32 biomarkers (28 blood and 4 urinary biomarkers) common to both studies, we compared gene-biomarker relationships that achieved significance (p<1×10^−8^) in the collapsing analysis with gene-biomarker relationships corresponding to the significant coding variant associations reported in the GWAS. We considered a comparatively relaxed significance threshold of p=1×10^−7^ for the GWAS results in order to be stringent when attributing a gene-biomarker relationship as being specific to the collapsing analysis.

We also hypothesised that the various variant-level “purifying” filters implemented for QV selection in the collapsing analysis can enable a more direct estimate for the effect of gene aberrations (e.g., PTVs) on biomarker levels. To investigate this hypothesis, we compared the effect sizes for gene-biomarker relationships that achieved significance in both the gene-level collapsing analysis and the microarray-based GWAS. For each such gene-biomarker relationship, we selected: (i) the *model* with the highest absolute beta in the collapsing analysis, and (ii) the individual *variant* with the highest absolute beta as reported in the Sinnott-Armstrong *et al* GWAS^14^. For the latter, we adopted the absolute beta estimated in the genotypic model in our ExWAS (for the corresponding gene-biomarker relationship) as a substitute, to account for possible differences in trait transformation, association model or covariates between our study and the Sinnott-Armstrong *et al* GWAS. Nonetheless, the absolute betas were highly correlated between the Sinnott-Armstrong *et al* GWAS and our ExWAS (Spearman’s rho=0.99) (**Figure S5**). We then compared the absolute beta of the collapsing model [step (i)] with that of the individual variant [step (ii)]. This approach provides a means to compare the effect size of aberrations in genes on biomarker levels estimated from individual coding variants captured by microarrays with that estimated from an aggregate of rare coding variants identified using exome sequencing.

### Estimating gene similarity based on association signatures from collapsing analysis

We aimed to leverage the rich catalogue of gene-level association statistics from the collapsing analysis – ascertained for the set of studied metabolic biomarkers and under different QV models – to identify genes that possess similar metabolic biomarker fingerprint as a (target) gene of interest. Such a ‘gene similarity’ metric can provide opportunities to not only expand our understanding of gene networks, but also offer alternative candidates in cases of difficult-to-drug targets. Gene-SCOUT (Gene Similarity from Continuous Traits)^24^, the tool that we developed for this purpose, can also estimate “similarity” between genes based on any set of quantitative traits of interest.

Rather than calculating similarities between genes directly, Gene-SCOUT estimates distances between genes, which it then uses as a proxy for their similarity. Based on that, the set of genes having the smallest distance from a given seed gene represent those that are most ‘similar’ to it. We applied the cosine distance method – which is commonly used in natural language processing^37^ – to calculate distances between genes^38^ based on their effects on the metabolic biomarkers (referred to as ‘features’) estimated in the collapsing analysis. In order to minimise the impact of stochastic effects on the gene similarity estimations, for a given seed gene of interest, only those features that the genes is associated with at p<1×10^−5^ are selected (‘feature selection’ step), guided by sensitivity analyses performed for a range of p-value thresholds^24^. Thus, distances from genes having p>1×10^−5^ for all features in common with the seed gene are not considered.

The feature set used to generate the Gene-SCOUT results comprised of the 198 metabolic blood biomarkers. Though there is a degree of correlation in our feature set (**Figure 2a**), we have demonstrated through simulations that correlation between features has minimal impact on gene similarity estimations^24^.

To illustrate the tool’s utility, we generated a network figure showing the genes that were most similar to each of the 24 genes that were significantly associated with LDL-cholesterol in the collapsing analysis.

## Data Availability

All data produced in the present study are available upon reasonable request to the authors

## Ethics Reporting

The protocols for UKB are overseen by The UK Biobank Ethics Advisory Committee (EAC); for more information see: https://www.ukbiobank.ac.uk/ethics/ and https://www.ukbiobank.ac.uk/wp-content/uploads/2011/05/EGF20082.pdf.

## Acknowledgements

We thank the participants and investigators in the UKB study who made this work possible (Resource Application Number 26041); the UKB Exome Sequencing Consortium (UKB-ESC) members AbbVie, Alnylam Pharmaceuticals, AstraZeneca, Biogen, Bristol-Myers Squibb, Pfizer, Regeneron and Takeda for funding the generation of the exome sequence data; the Regeneron Genetics Center for completing the sequencing and initial quality control of the exome sequencing data; and the AstraZeneca Centre for Genomics Research Analytics and Informatics team for processing and analysis of sequencing data.

## Author Contributions

S.P. designed the study. A.N., L.M., R.S.D., D.V., E.W., Q.W. and S.P. performed the analyses and statistical interpretation. A.N., R.S.D., A.R.H. and S.P. drafted the manuscript. All authors contributed to the review and critical revision of the manuscript.

## References

1. Willer, C. J. et al. Discovery and refinement of loci associated with lipid levels. Nature genetics 45, (2013).

2. Wuttke, M. et al. A catalog of genetic loci associated with kidney function from analyses of a million individuals. Nature genetics 51, (2019).

3. Kettunen, J. et al. Genome-wide association study identifies multiple loci influencing human serum metabolite levels. Nature genetics 44, (2012).

4. Yet, I. et al. Genetic Influences on Metabolite Levels: A Comparison across Metabolomic Platforms. PloS one 11, (2016).

5. Suhre, K. et al. A genome-wide association study of metabolic traits in human urine. Nature genetics 43, (2011).

6. Shin, S.-Y. et al. An atlas of genetic influences on human blood metabolites. Nature genetics 46, (2014).

7. Klarin, D. et al. Genetics of blood lipids among ∼300,000 multi-ethnic participants of the Million Veteran Program. Nature genetics 50, (2018).

8. Chambers, J. C. et al. Genome-wide association study identifies loci influencing concentrations of liver enzymes in plasma. Nature genetics 43, (2011).

9. Prins, B. P. et al. Genome-wide analysis of health-related biomarkers in the UK Household Longitudinal Study reveals novel associations. Scientific reports 7, (2017).

10. Wheeler, E. et al. Impact of common genetic determinants of Hemoglobin A1c on type 2 diabetes risk and diagnosis in ancestrally diverse populations: A transethnic genome-wide meta-analysis. PLoS medicine 14, (2017).

11. Long, T. et al. Whole-genome sequencing identifies common-to-rare variants associated with human blood metabolites. Nature genetics 49, (2017).

12. Gallagher, M. D. & Chen-Plotkin, A.S. The Post-GWAS Era: From Association to Function. American journal of human genetics 102, (2018).

13. Bycroft, C. et al. The UK Biobank resource with deep phenotyping and genomic data. Nature 562, 203–209 (2018).

14. Sinnott-Armstrong, N. et al. Genetics of 35 blood and urine biomarkers in the UK Biobank. Nature genetics 53, (2021).

15. UK10K Consortium et al. The UK10K project identifies rare variants in health and disease. Nature 526, (2015).

16. MacArthur, D. G. et al. A systematic survey of loss-of-function variants in human protein-coding genes. Science (New York, N.Y.) 335, (2012).

17. Ritchie, S. C. et al. Quality control and removal of technical variation of NMR metabolic biomarker data in ∼120,000 UK Biobank participants. medRxiv 2021.09.24.21264079 (2021) doi:10.1101/2021.09.24.21264079.

18. Gandotra, S. et al. Perilipin deficiency and autosomal dominant partial lipodystrophy. The New England journal of medicine 364, (2011).

19. Shin, G.-C., Kang, H. S., Lee, A. R. & Kim, K.-H. Hepatitis B virus-triggered autophagy targets TNFRSF10B/death receptor 5 for degradation to limit TNFSF10/TRAIL response. Autophagy 12, (2016).

20. Wang, Q. et al. Rare variant contribution to human disease in 281,104 UK Biobank exomes. Nature (2021) doi:10.1038/s41586-021-03855-y.

21. Aimee M. Deaton et al. Gene-level analysis of rare variants in 379,066 whole exome sequences identifies an association of GIGYF1 loss of function with type 2 diabetes. Scientific Reports 11, (2021).

22. Jurgens, S. J. et al. Rare Genetic Variation Underlying Human Diseases and Traits: Results from 200,000 Individuals in the UK Biobank. bioRxiv 2020.11.29.402495 (2020) doi:10.1101/2020.11.29.402495.

23. Chen, J. et al. The trans-ancestral genomic architecture of glycemic traits. Nature genetics 53, (2021).

24. Lawrence Middleton et al. Gene-SCOUT: identifying genes with similar continuous trait fingerprints from phenome-wide association analyses. Nucleic Acids Res (in submission) (2021).

25. Cohen, J. C., Boerwinkle, E., Mosley, T. H. & Hobbs, H. H. Sequence variations in PCSK9, low LDL, and protection against coronary heart disease. The New England journal of medicine 354, (2006).

26. Abul-Husn, N. S. et al. A Protein-Truncating HSD17B13 Variant and Protection from Chronic Liver Disease. The New England journal of medicine 378, (2018).

27. Akbari, P. et al. Sequencing of 640,000 exomes identifies GPR75 variants associated with protection from obesity. Science (New York, N.Y.) 373, (2021).

28. Nag, A. et al. Human genetic evidence supports MAP3K15 inhibition as a therapeutic strategy for diabetes. medRxiv 2021.11.14.21266328 (2021) doi:10.1101/2021.11.14.21266328.

29. Szustakowski, J. D. et al. Advancing human genetics research and drug discovery through exome sequencing of the UK Biobank. Nature genetics 53, (2021).

30. Cingolani, P. et al. A program for annotating and predicting the effects of single nucleotide polymorphisms, SnpEff: SNPs in the genome of Drosophila melanogaster strain w1118; iso-2; iso-3. Fly 6, 80–92 (2012).

31. Traynelis, J. et al. Optimizing genomic medicine in epilepsy through a gene-customized approach to missense variant interpretation. Genome research 27, 1715– 1729 (2017).

32. Ioannidis, N. M. et al. REVEL: An Ensemble Method for Predicting the Pathogenicity of Rare Missense Variants. American journal of human genetics 99, 877–885 (2016).

33. Manichaikul, A. et al. Robust relationship inference in genome-wide association studies. Bioinformatics 26, (2010).

34. Millard, L. A. C., Davies, N. M., Gaunt, T. R., Davey Smith, G. & Tilling, K. Software Application Profile: PHESANT: a tool for performing automated phenome scans in UK Biobank. International journal of epidemiology 47, (2018).

35. Fadista, J., Manning, A. K., Florez, J. C. & Groop, L. The (in)famous GWAS P-value threshold revisited and updated for low-frequency variants. European journal of human genetics : EJHG 24, (2016).

36. Petrovski, S. et al. An Exome Sequencing Study to Assess the Role of Rare Genetic Variation in Pulmonary Fibrosis. American journal of respiratory and critical care medicine 196, (2017).

37. Huang A. Similarity Measures for Text Document Clustering. NZCSRSC (2008).

38. Kittipong Chomboon, Pasapitch Chujai, Pongsakorn Teerarassamee, Kittisak Kerdprasop & Nittaya Kerdprasop. An Empirical Study of Distance Metrics for k-Nearest Neighbor Algorithm. Proceedings of the 3rd International Conference on Industrial Application Engineering (2015).

