## Supplemental Information for "Assessing the contribution of rare-to-common protein-coding variants to circulating metabolic biomarker levels via 412,394 UK Biobank exome sequences"

*Supplementary Information*

Abhishek Nag^1^, Lawrence Middleton^1^, Ryan S. Dhindsa^2,3^, Dimitrios Vitsios^1^, Eleanor Wigmore^1^, Erik L. Allman^1^, Anna Reznichenko^4^, Keren Carss^1^, Katherine R. Smith^1^, Quanli Wang^2^, Benjamin Challis^4^, Dirk S. Paul^1^, Andrew R. Harper^1^, Slavé Petrovski^1^

^1^Centre for Genomics Research, Discovery Sciences, BioPharmaceuticals R&D, AstraZeneca, Cambridge, UK

^2^Centre for Genomics Research, Discovery Sciences, BioPharmaceuticals R&D, AstraZeneca, Waltham, USA

^3^Department of Molecular and Human Genetics, Baylor College of Medicine and Jan and Dan Duncan Neurological Research Institute at Texas Children's Hospital, Houston, TX 77030, USA

^4^Translational Science and Experimental Medicine, Early CVRM, BioPharmaceuticals R&D, AstraZeneca, Gothenburg, Sweden

**Corresponding author:**
Slavé Petrovski

Vice-President, Centre for Genomics Research,

Discovery Sciences, BioPharmaceuticals R&D
AstraZeneca
Cambridge

United Kingdom


**Supplementary Figures**

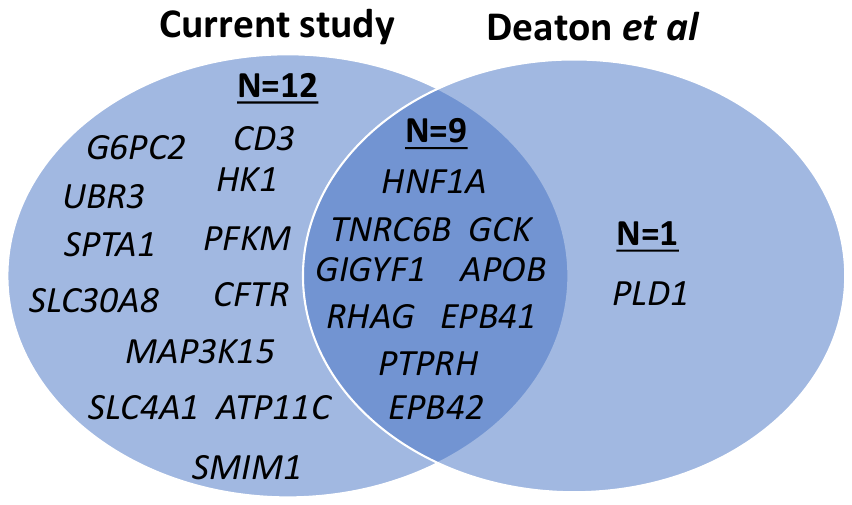

**Figure S1: Comparison of the genes for which PTV-based signals were identified for HbA1c in our gene-level collapsing analysis and by Deaton *et al***^21^

A gene-level collapsing analysis was conducted by Deaton *et al*^21^ in the same UKB exome sequencing dataset described in this study but applying a different analysis framework and model parameters. A comparison of the genes for which PTV-based signals were observed for HbA1c (adopting Deaton *et al*’s significance cut-off of p<8x10^-7^) in the two analyses is shown.

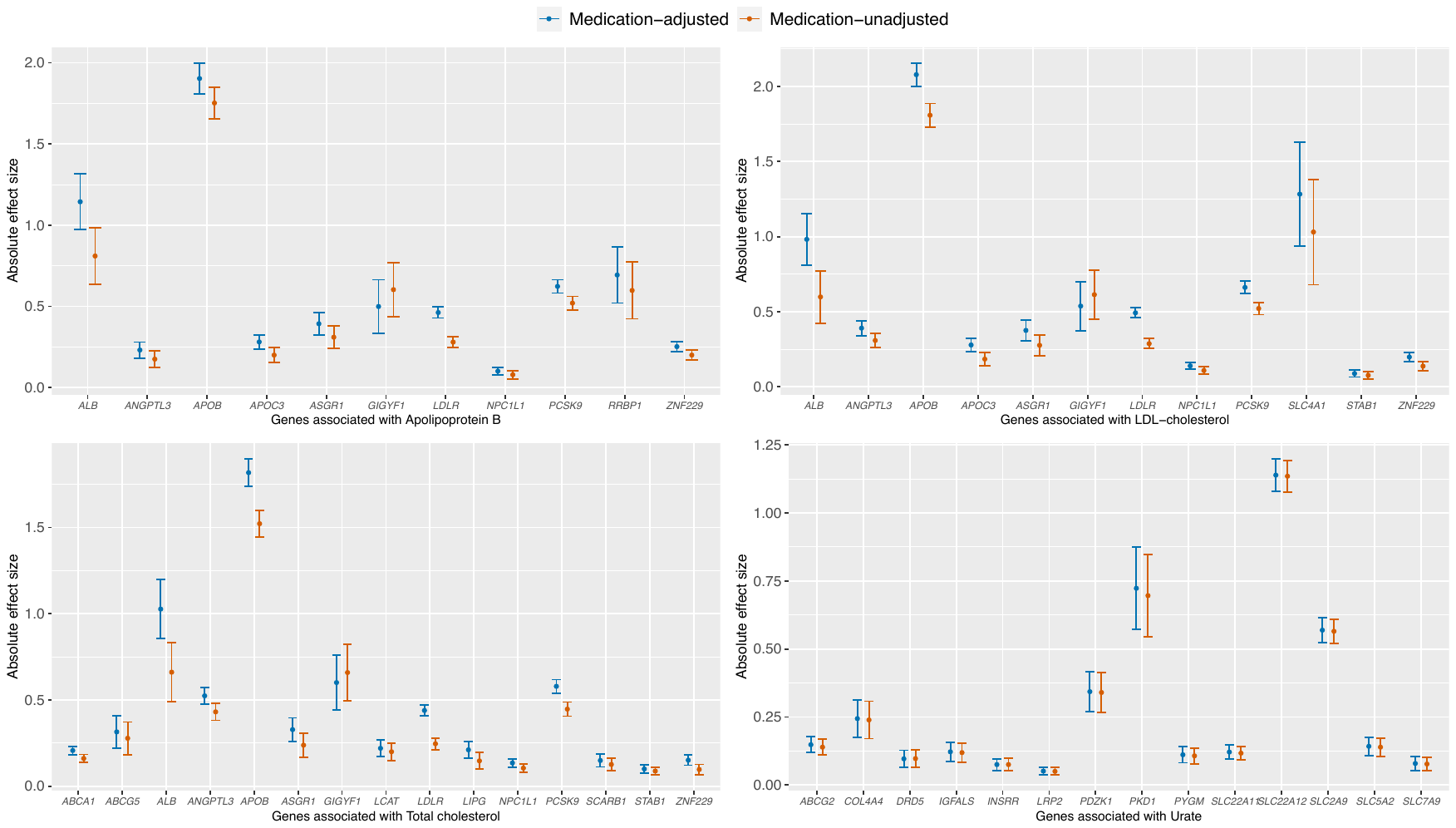

**Figure S2: Comparing the effect sizes for the gene-biomarker relationships that were significantly associated in both the medication-unadjusted and the medication-adjusted collapsing analyses**
Gene-level collapsing analysis was performed for the medication-adjusted values for apolipoprotein B, LDL-cholesterol and total cholesterol (all adjusted for the effect of statins), and urate (adjusted for the effect of allopurinol). For each of these four biomarkers, we compared the effect sizes between the medication-unadjusted and the medication-adjusted collapsing analyses for the genes that were significantly associated in both.

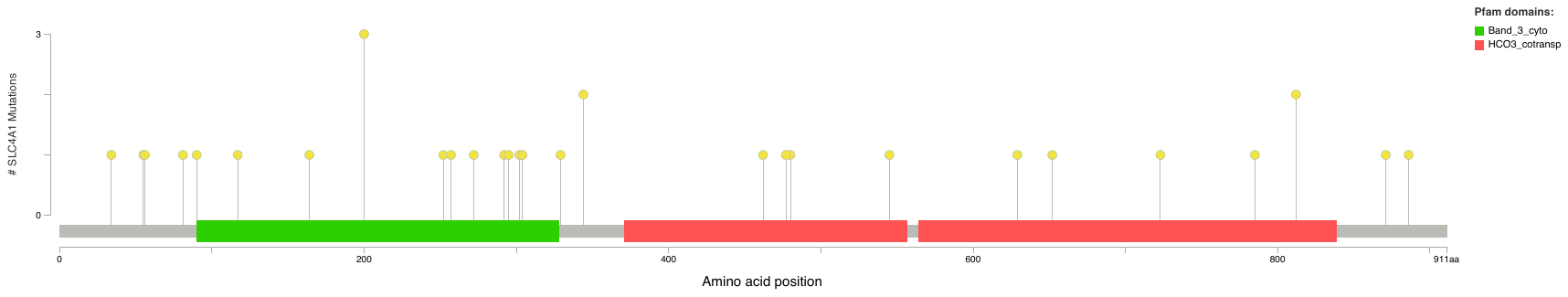

**Figure S3: Location of the protein-truncating variants (PTVs) in the *SLC4A1* gene sequence**

Significant associations were observed between PTVs in *SLC4A1* (“ptv” model) and bilirubin (total and direct), LDL-cholesterol and HbA1c in the gene-level collapsing analysis. The figure shows the location and the number of carriers for PTVs in the *SLC4A1* gene sequence ­– as noted, 25/28 PTVs had just a single carrier, demonstrating the high allelic heterogeneity that underlies the associations observed for this ge

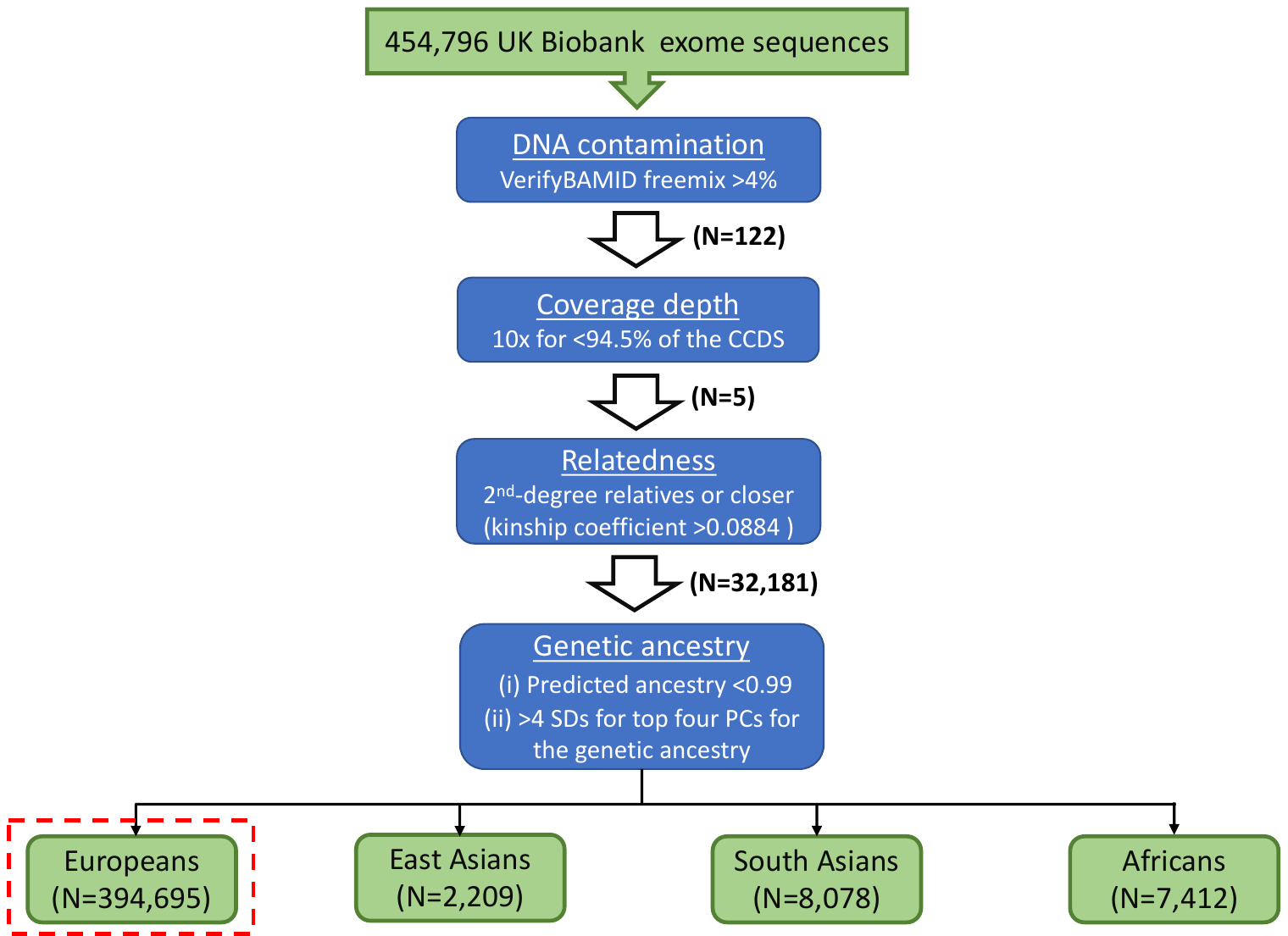

**Figure S4: Flowchart of the sample-level quality control measures applied to the UK Biobank exome sequences**

The figure illustrates the various quality control measures that were applied for excluding exome sequencing samples from the UK Biobank before carrying them forward for the association analyses.

The number of samples that were excluded at each stage of quality control have been provided in the parentheses.
(CCDS = consensus coding sequence; PC = principal component; SD = standard deviation)

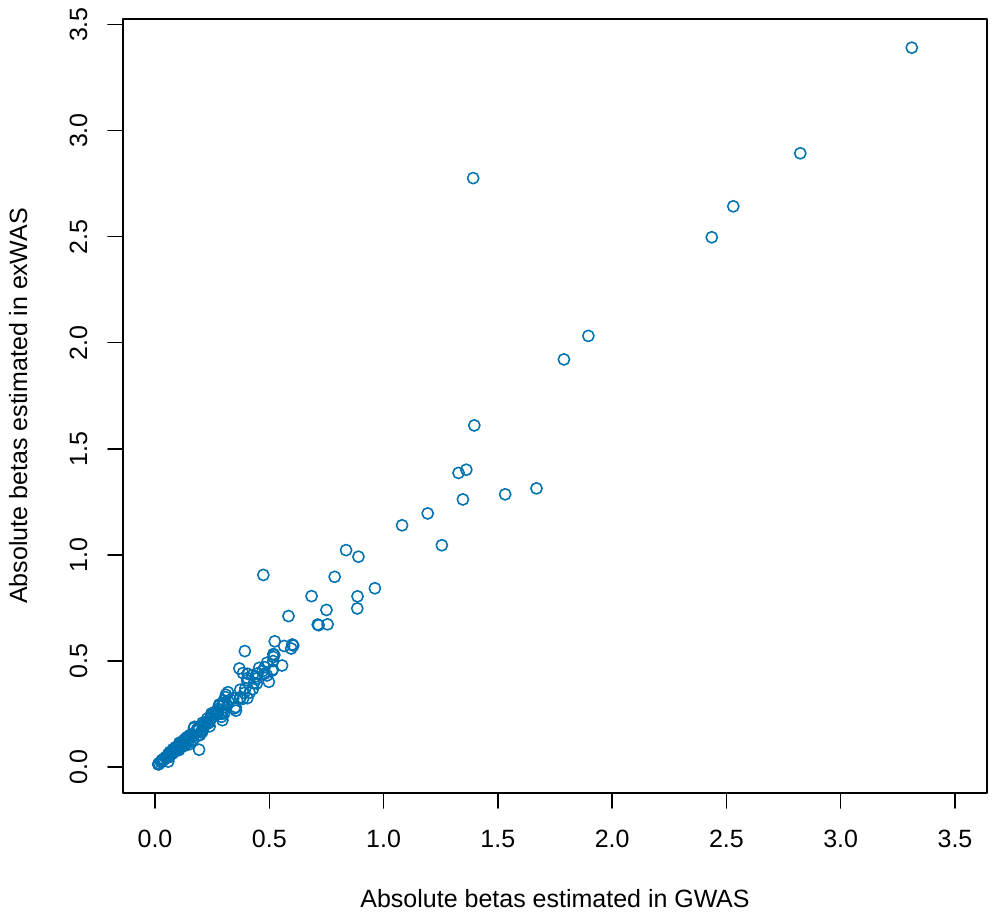
**Figure S5: Correlation between the absolute effect sizes estimated in the ExWAS and the microarray-based GWAS**

For the gene-biomarker relationships that were significantly associated in both the collapsing analysis and the Sinnott-Armstrong *et al* microarray-based GWAS^14^ (N=215), the effect size for the variants with the highest absolute effect size in the microarray-based GWAS was highly correlated with that estimated in the ExWAS.

**Supplementary Tables**

**Tables S1A and 1B: Summary of the metabolic blood biomarkers from the UK Biobank that were analysed in this study**

Provided as separate additional file

The 198 metabolic blood biomarkers [30 clinical biomarkers (Table S1A) and the 168 metabolites (Table S1B)] from the UK Biobank that were analysed in this study have been summarised.

Rank-based inverse-normal transformation was applied to each metabolic biomarker prior to performing the association analyses.

**Tables S2A and S2B: Distinct relationships between genes and metabolic biomarkers identified in the single variant analysis (ExWAS) in the European ancestry participants**

Provided as separate additional file

The tables summarise the significant relationships (p<1x10^-8^) between genes and metabolic biomarkers identified in the single variant analysis for the 30 clinical biomarkers (Table S2A) and the 168 metabolites (Table S2B) in the European ancestry participants. For each significant gene-biomarker relationship, the most significant variant and genetic model, along with the corresponding association statistics have been provided.

‘NA’s are used to indicate association statistics for biomarkers for which medication-adjusted analysis was not performed.
*Single variant analysis for the medication-adjusted values was conducted for apolipoprotein B, LDL-cholesterol and total cholesterol (all adjusted for statins), and for urate (adjusted for allopurinol).
(Chr = Chromosome; SE = Standard Error; MAF = Minor Allele Frequency)

**Tables S3A and S3B: Significant protein truncating variant (PTV) associations identified for the metabolic blood biomarkers in the European ancestry participants**

Provided as separate additional file

The tables provide the significantly associated PTVs for the 30 clinical biomarkers (Table S3A) and the 168 metabolites (Table S3B) in the European ancestry participants. Of the total 1,366 significant PTV associations, 602 (44%) were attributable to rare PTVs (MAF<0.1%) with large effect sizes (>0.5 SD).
*When the PTV-biomarker association was significant in the medication-adjusted analysis only, the medication-adjusted association statistics have been provided.

(Chr = Chromosome; SE = Standard Error; MAF = Minor Allele Frequency)

**Tables S4A and S4B: Distinct relationships between genes and metabolic biomarkers identified in the gene-level collapsing analysis in the European ancestry participants**

Provided as separate additional file

The tables summarise the significant (p<1x10^-8^) relationships between genes and metabolic biomarkers identified in the gene-level collapsing analysis for the 30 clinical biomarkers (Table S4A) and the 168 metabolites (Table S4B) in the European ancestry participants. For each significant gene-biomarker relationship, the most significant collapsing model and the corresponding association statistics have been provided.
‘NA’s are used to indicate association statistics for biomarkers for which medication-adjusted analysis was not performed.

*Collapsing analysis for medication-adjusted values was conducted for apolipoprotein B, LDL-cholesterol and total cholesterol (all adjusted for statins), and for urate (adjusted for allopurinol).
(Chr = Chromosome; SE = Standard Error; QV = Qualifying Variants)

**Table S5A and S5B: Distinct relationships between genes and metabolic biomarkers identified in the gene-level collapsing analysis combining all ancestral groups**

Provided as separate additional file

The tables summarise the significant (p<1x10^-8^) relationships between genes and metabolic biomarkers identified in the gene-level collapsing analysis for the 30 clinical biomarkers (Table S5A) and the 168 metabolites (Table S5B) combining all ancestral groups. For each significant gene-biomarker relationship, the most significant collapsing model and the corresponding association statistics have been provided.

In addition to the 394,695 European ancestry participants in the UKB, 8,078 South Asians, 2,209 East Asians and 7,412 Africans were included in this analysis.
(Chr = Chromosome; SE = Standard Error; QV = Qualifying Variants)

**Table S6: Collapsing models that captured the gene-biomarker relationships that were specific to the collapsing analysis (i.e., not detected in ExWAS)**

The gene-biomarker relationships that were specific to the collapsing analysis (i.e., not detected in ExWAS) were stratified based on the different types of collapsing models / MAF bins.

**Table S7: Distinct gene-biomarker associations identified in the gene-level collapsing analysis for the urinary biomarkers**

Provided as separate additional file

The table summarises the significant (p<1x10^-8^) gene-biomarker relationships identified in the gene-level collapsing analysis for the four urinary biomarkers. For each gene-biomarker relationship, the most significant collapsing model and the corresponding association statistics have been provided.
(Chr = Chromosome; SE = Standard Error; QV = Qualifying Variants)

**Table S8: Gene-biomarker relationships from the gene-level collapsing analysis that were not detected in the microarray-based GWAS**

Provided as separate additional file

The significant gene-biomarker relationships (p<1x10^-8^) identified in the gene-level collapsing analysis were compared with that reported in the microarray-based GWAS (p<1x10^-7^) for the 28 blood and 4 urinary biomarkers that were analysed in both studies. This table provides the list of gene-biomarker relationships that were specific to the collapsing analysis i.e., not detected in the microarray-based GWAS.
(Chr = Chromosome; SE = Standard Error)

**Table S9: Comparison of effect size estimates between the gene-level collapsing analysis and the microarray-based GWAS**

Provided as separate additional file

A comparison of the effect sizes for the gene-biomarker relationships that were significantly associated in both the gene-level collapsing analysis and the Sinnott-Armstrong *et al* microarray-based GWAS^14^ was performed (N=215). For each such gene-biomarker relationship, the collapsing model with the highest absolute beta in the collapsing analysis and the individual variant having the highest absolute beta in the microarray-based analysis was selected. For the latter, we used the corresponding absolute beta estimated in our ExWAS (‘genotypic’ model) as a substitute, to account for possible differences (such as trait transformation, covariates or association model used) between our study and the microarray-based GWAS.

**Table S10: Effect of PTVs in *SLC4A1* on direct bilirubin, HbA1c and LDL-cholesterol adjusted for hereditary spherocytosis and haemolytic anaemia**

The associations for PTVs in *SLC4A1* (“ptv” model) with direct bilirubin, HbA1c and LDL-cholesterol were adjusted for hereditary spherocytosis and haemolytic anaemia using both binary disease outcomes (ICD-10 code-based) and relevant quantitative red blood cell indices (mean corpuscular haemoglobin concentration and red cell distribution width).

**Table S11: Summary of the different models implemented in the gene-level collapsing analysis**

The table provides the criteria used for determining qualifying variants (QVs) for the 10 different models used in the gene-level collapsing analysis. In addition, a synonymous collapsing model was used for the purpose of establishing an empirical negative control.

**Table S6**

| **Type of collapsing models / MAF bins** | **Number of gene-biomarker relationships specific to the collapsing analysis (i.e., not detected in ExWAS) that were captured (out of 96) [%]** |
| --- | --- |
| **Only Ultra-rare**  [UR/URmtr] (MAF≤0.005%) | 15 [15.6%] |
| **Ultra-rare + rare**  [UR/URmtr/raredmg/raredmgmtr]  (MAF≤0.025%)) | 18 [18.8%] |
| **Ultra-rare + rare + PTV (rare)** [UR/URmtr/raredmg/raredmgmtr/ ptv/ptvraredmg] (MAF≤0.1%) | 78 [81.3%] |
| **Ultra-rare + rare + PTV (rare) + flex** [UR/URmtr/raredmg/raredmgmtr/ ptv/ptvraredmg/flexdmg/flexnonsynmtr] (MAF≤0.1%) | 93 [96.9%] |
| **Ultra-rare + rare + PTV (all) + flex + rec** [UR/URmtr/raredmg/raredmgmtr/ ptv/ptvraredmg/flexdmg/ flexnonsynmtr/ptv5pcnt/rec] (MAF≤5%) | 96 [100%] |

**Table S10**

| **Clinical biomarker** | ***SLC4A1* collapsing signals** | | ***SLC4A1* collapsing signals adjusted for binary disease outcomes** | | | | ***SLC4A1* collapsing signals adjusted for quantitative red blood cell indices** | | | |
| --- | --- | --- | --- | --- | --- | --- | --- | --- | --- | --- |
|  |  |  | **Hereditary spherocytosis** | | **Haemolytic anaemia** | | **Mean corpuscular haemoglobin concentration (MCHC)** | | **Red cell distribution width  (RDW)** | |
|  | **Beta** | **p-value** | **Beta** | **p-value** | **Beta** | **p-value** | **Beta** | **p-value** | **Beta** | **p-value** |
| **Direct bilirubin** | 2.01 | 1.84E-28 | 1.56 | 6.38E-17 | 1.85 | 3.31E-24 | 1.89 | 1.38E-24 | 2.02 | 6.94E-28 |
| **HbA1c** | -2.21 | 1.44E-25 | -1.85 | 4.89E-18 | -2.15 | 3.31E-24 | -1.97 | 5.14E-20 | -2.30 | 4.28E-27 |
| **LDL-cholesterol** | -1.03 | 8.00E-09 | -0.87 | 2.51E-06 | -0.91 | 4.58E-07 | -1.07 | 3.45E-09 | -0.99 | 4.83E-08 |

**Table S11**

| **Collapsing model** | **gnomAD  MAF*** | **UKB  MAF** | **UKB cohort  no call or QC fail^** | **Variant  type** | **REVEL**^32^  **cut-off** | **Missense tolerance ratio (MTR)**^31^ **cut-offs** |
| --- | --- | --- | --- | --- | --- | --- |
| **syn (synonymous negative control)** | ≤0.005% | ≤0.05% | ≤0.005% | Synonymous | - | - |
| **ptv      (Protein Truncating)** | ≤0.1% (popmax) | ≤0.1% | ≤0.01% | PTV | - | - |
| **ptv5pcnt (Protein Truncating, ≤5% MAF)** | ≤5% (popmax) | ≤5% | ≤0.5% | PTV | - | - |
| **UR      (Ultra-rare damaging)** | 0% | ≤0.005% | ≤0.001% | Non-synonymous | ≥0.25 | - |
| **URmtr (Ultra-rare damaging, MTR informed)** | 0% | ≤0.005% | ≤0.001% | Non-synonymous | ≥0.25 | MTR≤25^th^ %ile or intragenic MTR≤50^th^ %ile |
| **raredmg (Rare damaging)** | ≤0.005% | ≤0.025% | ≤0.005% | Missense | ≥0.25 | - |
| **raredmgmtr (Rare damaging, MTR informed)** | ≤0.005% | ≤0.025% | ≤0.005% | Missense | ≥0.25 | MTR≤25^th^ %ile or  intragenic MTR≤50^th^ %ile |
| **flexdmg (Flexible MAF, damaging non-synonymous)** | ≤0.1% (popmax) | ≤0.1% | ≤0.01% | Non-synonymous | ≥0.25 | - |
| **flexnonsynmtr (Flexible MAF, non-synonymous, MTR informed)** | ≤0.1% (popmax) | ≤0.1% | ≤0.01% | Non-synonymous | - | MTR≤25^th^ %ile or  intragenic MTR≤50^th^ %ile |
| **ptvraredmg (PTV or rare damaging models combined)** | PTV≤0.1% (popmax)  missense≤0.005%  and ≤0.05% (popmax) | PTV≤0.1%, missense≤ 0.025% | ≤0.01% | Non-synonymous | ≥0.25 | - |
| **rec (Non-synonymous recessive)** | ≤1% (popmax)  ≤10 homozygous calls | ≤1% | ≤0.1% | Non-synonymous | - | - |

(MAF = minor allele frequency; QC = quality control; MTR = Missense Tolerance Ratio)
*reflects the gnomAD global_raw MAF unless otherwise specified. 
^reflects the maximum proportion of UKB exome sequences permitted to either have ≤ 10-fold coverage at variant site or carry a low-confidence variant that did not meet one of the quality-control thresholds applied to collapsing analyses (see methods). 
**Synonymous**: synonymous_variant 
**PTV**: exon_loss_variant, frameshift_variant, start_lost, stop_gained, stop_lost, splice_acceptor_variant, 
splice_donor_variant, gene_fusion, bidirectional_gene_fusion, rare_amino_acid_variant, transcript_ablation 
**Missense**: missense_variant_splice_region_variant, missense_variant 
**Nonsynonymous**:exon_loss_variant, frameshift_variant, start_lost, stop_gained, stop_lost, splice_acceptor_variant,splice_donor_variant, gene_fusion, bidirectional_gene_fusion, rare_amino_acid_variant, transcript_ablation, 
conservative_inframe_deletion, conservative_inframe_insertion, disruptive_inframe_insertion, 
disruptive_inframe_deletion, missense_variant_splice_region_variant, missense_variant, protein_altering_variant
